# Ensemble of deep masked language models for effective named entity recognition in multi-domain corpora

**DOI:** 10.1101/2021.04.26.21256038

**Authors:** Nona Naderi, Julien Knafou, Jenny Copara, Patrick Ruch, Douglas Teodoro

## Abstract

The health and life science domains are well known for their wealth of entities. These entities are presented as free text in large corpora, such as biomedical scientific and electronic health records. To enable the secondary use of these corpora and unlock their value, named entity recognition (NER) methods are proposed. Inspired by the success of deep masked language models, we present an ensemble approach for NER using these models. Results show statistically significant improvement of the ensemble models over baselines based on individual models in multiple domains - chemical, clinical and wet lab - and languages - English and French. The ensemble model achieves an overall performance of 79.2% macro F_1_-score, a 4.6 percentage point increase upon the baseline in multiple domains and languages. These results suggests that ensembles are a more effective strategy for tackling NER. We further perform a detailed analysis of their performance based on a set of entity properties.

## 1 Introduction

In the health and life science domains, most of the information is encoded in free text reports. For example, it is estimated that around 90% of electronic health records (EHR) data is available as free text. While text format facilitates capturing information, it makes the secondary use of the data challenging. To support data structuring and unlock the value of textual databases in secondary usage applications, named entity recognition (NER) methods have been proposed. NER is the task of detecting entities in text and assigning concept names, or categories, to them. The health and life science domains (e.g., biology, chemistry, medicine, etc.) are notoriously known for the wealth of named entities and synonyms, for example, microorganism taxonomies, drug brands, and gene names, to name a few. This richness of named entities together with the variety of formats and (mis)spellings make NER in health and life sciences corpora (e.g., EHR, lab protocols, scientific publications, patents, etc.) a challenging task.

Traditional NER methods mainly involve dictionary and rule-based, machine learning, and hybrid methods that combine these approaches [Quimbaya et al., 2016]. They normally require domain knowledge and feature engineering. Fully supervised NER approaches include Support Vector Machines (SVM), decision trees, hidden Markov models [Zhao, 2004], and conditional fields (CRFs) [Li et al., 2008, Leaman et al., 2015, Rocktäschel et al., 2012]. They are often used to provide a baseline for model evaluation.

More recently, deep masked language models trained on large corpus have achieved state-of-the-art in most NLP related tasks, including NER. Bidirectional Encoder Representations from Transformers (BERT) [Devlin et al., 2019] was the first to explore the transformer architecture as a general framework for NLP [Vaswani et al., 2017]. Once the model is trained (or pre-trained in the BERTology parlance) on a large corpus, it can be adapted and effectively used in specialised downstream NLP tasks, such as question-answering, text classification and NER, leveraging the feature representations learned by the model during the pre-training phase.

Since the advent of BERT, a myriad of transformer-based masked language models have been proposed [Liu et al., 2019, Yang et al., 2019, Alsentzer et al., 2019]. These models vary mostly in the tokenization used, how the masking is performed, and the trained data used during the pre-training phase. In this paper, we assess BERT-like models for NER in cross-domain (chemistry, clinical and wet lab) and -language (English and French) corpora. Particularly, we leverage different pre-trained models available in the literature to create ensembles of named entity recognizers. We evaluate our models in chemistry, clinical and wet lab corpora provided in the context of the ChEMU (Cheminformatics Elsevier Melbourne University) [He et al., 2020a], DEFT (Défi Fouille de Textes) [Grabar et al., 2018] and WNUT (Workshop on Noisy User-generated Text) [Tabassum et al., 2020] challenges, respectively. Our results show that ensembles of named entity recognizers based on masked language models can achieve effective NER performances in these different domains and languages. We further perform an analysis of certain entity properties, including entity lengths, entity frequencies, and consistencies for better understanding the performance of these models.

## 2 Related work

Deep learning approaches trained on large unstructured data have shown considerable success in NLP problems, including NER [Devlin et al., 2019, Liu et al., 2019, Lample et al., 2016, Beltagy et al., 2019, Jin et al., 2019]. These models use the learned representations over the large data and reuse them in a supervised setting for a downstream task. For domain-specific tasks, the models that are trained on large general text can be further trained on domain specific large data and then adapted for a downstream task [Lee et al., 2019, Gururangan et al., 2020, Alsentzer et al., 2019] or the models can be trained only on domain-specific data and then adapted for a specific task [Beltagy et al., 2019]. Furthermore, several models are proposed for cross-domain NER; however, only a few focus on biomedical or clinical NER [Jia et al., 2019]. One study is that of [Lee et al., 2018] that utilizes the idea of transfer learning to identify the named entities in i2b2 2014/2016 using the MIMIC dataset.

### 2.1 Chemical NER

To further improve the performance of the traditional approaches using hand-crafted features [Leaman et al., 2015, Rocktäschel et al., 2012, Habibi et al., 2016, Zhang et al., 2016, Akhondi et al., 2016], a number of studies leveraged the power of word embeddings in addition to the hand-crafted features in a single model (LSTM-CRF) [Habibi et al., 2017, Corbett and Boyle, 2018, Zhai et al., 2019, Hemati and Mehler, 2019]. These methods have shown a significant improvement over the traditional methods.

### 2.2 Clinical NER

Various NER challanges and shared tasks [Uzuner et al., 2010, Kelly et al., 2014, Névéol et al., 2015, Suominen et al., 2013, Bethard et al., 2015] fostered the development of NER methods Van Mulligen et al., 2016 Kim et al., 2015 Jiang et al., 2011, De Bruijn et al., 2011, El Boukkouri et al., 2019] for clinical domain in different languages [Lopes et al., 2019, Schneider et al., 2020, Sun and Yang, 2019]. Performance varies greatly across the different methods and corpora, with more modern methods achieving F_1_-score as high as 95%.

### 2.3 Wet lab NER

Luan et al. [2019] introduce a model based on dynamic span graph to jointly extract named entities and relations on wet lab protocols and other corpora. Wadden et al. [2019] build upon Luan et al. [2019]’s model by combining BERT and dynamic span graph. Dai et al. [2019] compute the similarity of the pre-train data and the data of the target application to investigate the effectiveness of pre-trained word vectors. They found that the effectiveness of the pre-trained word vectors depends on the vocabulary overlap of the source and target domains.

## 3 Data

In this section, we present the datasets used to assess the ensembles of masked languages models for the extraction of named entities in chemical, clinical and wet lab domains. The first dataset, provided in the context of the ChEMU 2020 challenge, consists of a collection of English chemistry patents annotated with chemical reaction entities. The second dataset, provided in the context of the DEFT 2020 challenge, consists of a collection of French EHR notes annotated clinical entities. Finally, the third dataset, provided in the context of the WNUT 2020 challenge, consists of English laboratory protocols annotated with wet lab entities.

### 3.1 Benchmark data for chemical entity recognition - ChEMU 2020 dataset

The ChEMU 2020 benchmark dataset^1^ [He et al., 2021] contains snippets sampled from 170 English patents from the European Patent Office and the United States Patent and Trademark Office [He et al., 2020a,b, 2021]. As shown in Fig. 1, these snippets are annotated with several chemical reaction entities, including *reaction_product, starting_material* and *temperature*. The training and test set of the ChEMU dataset contains a total of 1500 snippets (training: 1,125; test: 375) annotated with 26,857 entities (training: 20,186; test: 6,671) using the BRAT standoff format [Stenetorp et al., 2012].

**Figure 1:**
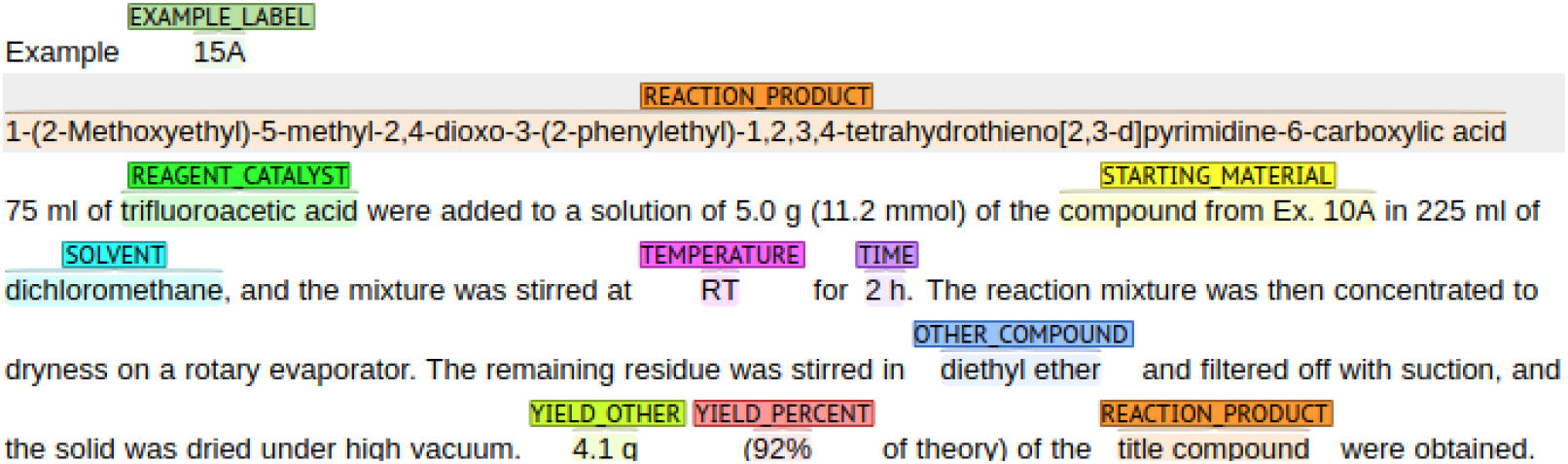
An example of a patent passage of the ChEMU dataset with entity annotations. The annotations are color coded, representing the different entities in the dataset

Table 1 shows the entity distribution for the training and test sets. The majority of the annotations are provided for the *other_compound, reaction_product* and *starting_material* entities, covering 52% of the examples in the training and test datasets. In contrast, *example_label, yield_other* and *yield_percent* entities represent together only 18% of entities in the training and test sets.

**Table 1:** Entity distribution in the training and test sets of ChEMU benchmark dataset.

| Entity | Train |  | Test |  |
| --- | --- | --- | --- | --- |
|  | Count | % | Count | % |
| example_label | 1,104 | 5.5 | 349 | 5.2 |
| other_compound | 5,720 | 28.3 | 1931 | 28.9 |
| reaction_product | 2,558 | 12.7 | 855 | 12.8 |
| reagent_catalyst | 1,570 | 7.8 | 504 | 7.6 |
| solvent | 1,390 | 6.9 | 428 | 6.4 |
| starting_material | 2,167 | 10.7 | 711 | 10.7 |
| temperature | 1,861 | 9.2 | 612 | 9.2 |
| time | 1,311 | 6.5 | 452 | 6.8 |
| yield_other | 1,322 | 6.5 | 440 | 6.6 |
| yield_percent | 1,183 | 5.9 | 389 | 5.8 |
| Total | 20,186 | 100.0 | 6,671 | 100.0 |

### 3.2 Benchmark data for clinical entity recognition - DEFT 2020 dataset

The DEFT benchmark dataset^2^, a subset of the CAS corpus [Grabar et al., 2018], is composed of 100 French clinical documents (training: 90; test: 10) manually annotated with the 8,098 entities (training: 7,421; test: 677) in the following categories: *pathologie, sosy* (symptoms and signs), *anatomie, dose, examen, mode, moment, substance, traitement*, and *valeur*. An example of a clinical note annotation is shown in Figure 2. We can notice that nested entities appear often in the annotations.

**Figure 2:**
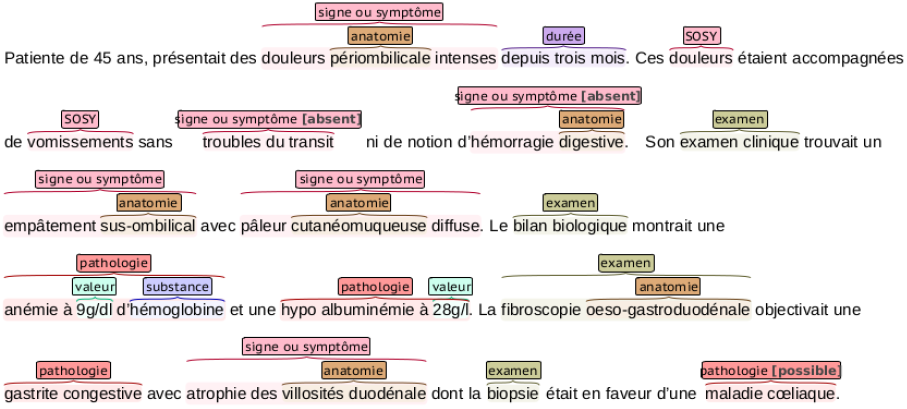
An example of a clinical narrative of the DEFT dataset with entity annotations. The annotations are color coded, representing the different entities in the dataset. Notice that some entities are nested.

Table 2 shows the distribution of annotations among the entities in the training and test datasets. The majority of annotations come from the *sosy, anatomie* and *examen* entities, which compose together 54% of the training data. On the other hand, *mode, dose* and *pathologie* represent together only 13% of the training dataset. In contrast to the ChEMU data, the distribution of the training and test sets vary significantly.

**Table 2:** Entity distribution in the training and test sets of the DEFT benchmark dataset.

| Entity | Train |  | Test |  |
| --- | --- | --- | --- | --- |
|  | Count | % | Count | % |
| anatomie | 1,298 | 17.5 | 174 | 25.7 |
| dose | 342 | 4.6 | 5 | 0.7 |
| examen | 1,081 | 14.6 | 137 | 20.2 |
| mode | 238 | 3.2 | 11 | 1.6 |
| moment | 440 | 5.9 | 54 | 8.0 |
| pathologie | 351 | 4.7 | 184 | 27.2 |
| sosy | 1,647 | 22.2 | 33 | 4.9 |
| substance | 968 | 13.0 | 22 | 3.3 |
| traitement | 494 | 6.7 | 52 | 7.7 |
| valeur | 562 | 7.6 | 5 | 0.7 |
| Total | 7,421 | 100.0 | 677 | 100.0 |

### 3.3 Benchmark data for wet lab entity recognition - WNUT 2020 dataset

The WNUT benchmark dataset [Kulkarni et al., 2018] is composed of 727 unique (English) wet lab protocols^3^ that describe experimental procedures. This dataset (training: 616; test: 111) was manually annotated with the 102,957 entities (training: 79,757; test: 23,200) in the following categories: *Action, Amount, Concentration, Device, Generic-Measure, Location, Measure-Type, Mention, Modifier, Numerical, Reagent, Seal, Size, Speed, Temperature, Time*, and *pH*. An example of a lab protocol annotation is shown in Figure 3.

**Figure 3:**
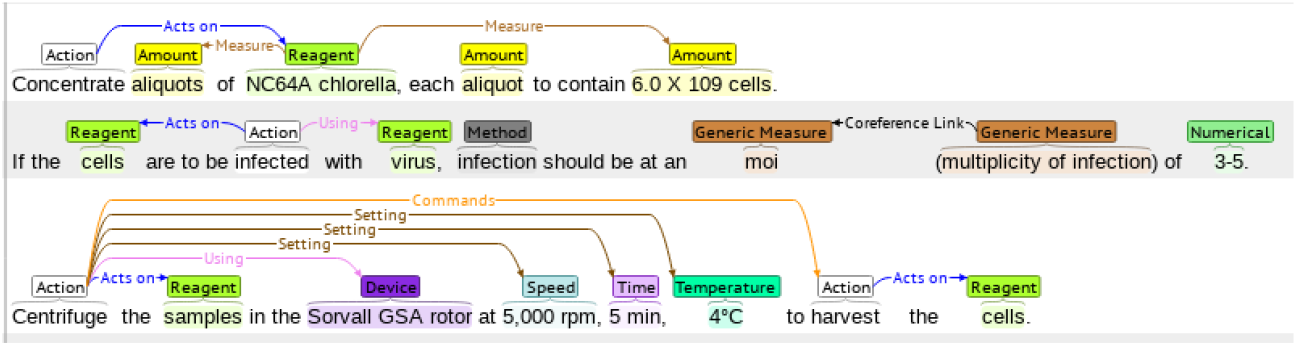
An example of a wet lab protocol of the WNUT dataset with entity annotations. The annotations are color coded, representing the different entities in the dataset.

In Table 3, we see the distribution of the 18 entities by each subset. As in the other chemical and clinical datasets, there is a significant class imbalance, with only two of them (*Action* and *Reagent*) representing more than 50% of annotations in the training set. This table also shows that, similar to the ChEMU dataset, the proportions of entities are fairly similar across the training and test subsets.

**Table 3:** Entity distribution in the training and test sets of the WNUT benchmark dataset.

| Entity | Train |  | Test |  |
| --- | --- | --- | --- | --- |
|  | Count | % | Count | % |
| Action | 20,504 | 25.7 | 5,346 | 23.0 |
| Amount | 5,712 | 7.2 | 1,223 | 5.3 |
| Concentration | 2,287 | 2.9 | 701 | 3.0 |
| Device | 2,836 | 3.6 | 888 | 3.8 |
| Generic-Measure | 759 | 1.0 | 173 | 0.8 |
| Location | 6,643 | 8.3 | 1,657 | 7.1 |
| Measure-Type | 1,453 | 1.8 | 720 | 3.1 |
| Mention | 396 | 0.5 | 142 | 0.6 |
| Method | 2,716 | 3.4 | 1,059 | 4.6 |
| Modifier | 7,736 | 9.7 | 3,416 | 14.7 |
| Numerical | 1,322 | 1.7 | 513 | 2.2 |
| Reagent | 18,710 | 23.5 | 5,012 | 21.6 |
| Seal | 366 | 0.5 | 119 | 0.5 |
| Size | 498 | 0.6 | 232 | 1.0 |
| Speed | 1,032 | 1.3 | 238 | 1.0 |
| Temperature | 2,610 | 3.3 | 744 | 3.2 |
| Time | 4,011 | 5.0 | 951 | 4.1 |
| pH | 166 | 0.2 | 66 | 0.3 |
| Total | 79,757 | 100.0 | 23,200 | 100.0 |

## 4 Method

In this section, we describe our methodology to fine-tune a single deep masked language model to recognize named entities in the chemical, clinical and wet lab domains in English and French corpora. Then, we detail how these different fine-tuned language models were combined to provide an ensemble NER model.

### 4.1 Single deep masked language model for NER

To build the ensemble NER model, we fine-tuned different individual masked languages models based on the transformers architecture [Vaswani et al., 2017]. In the case of NER, masked language models are fine-tuned using a specialised training set - in our case, the chemical, clinical and wet lab annotated corpora - to classify tokens according to the named entity classes. As shown in Table 4, we assessed single language models based on or derived from the BERT architecture. BERT was originally pretrained on a large corpus of English text extracted from Book-Corpus [Zhu et al., 2015] and Wikipedia, with different number of attention heads for the base and large types (12 and 24 transformer layers and hidden representations of 768 and 1024 dimensions, respectively).

**Table 4:**
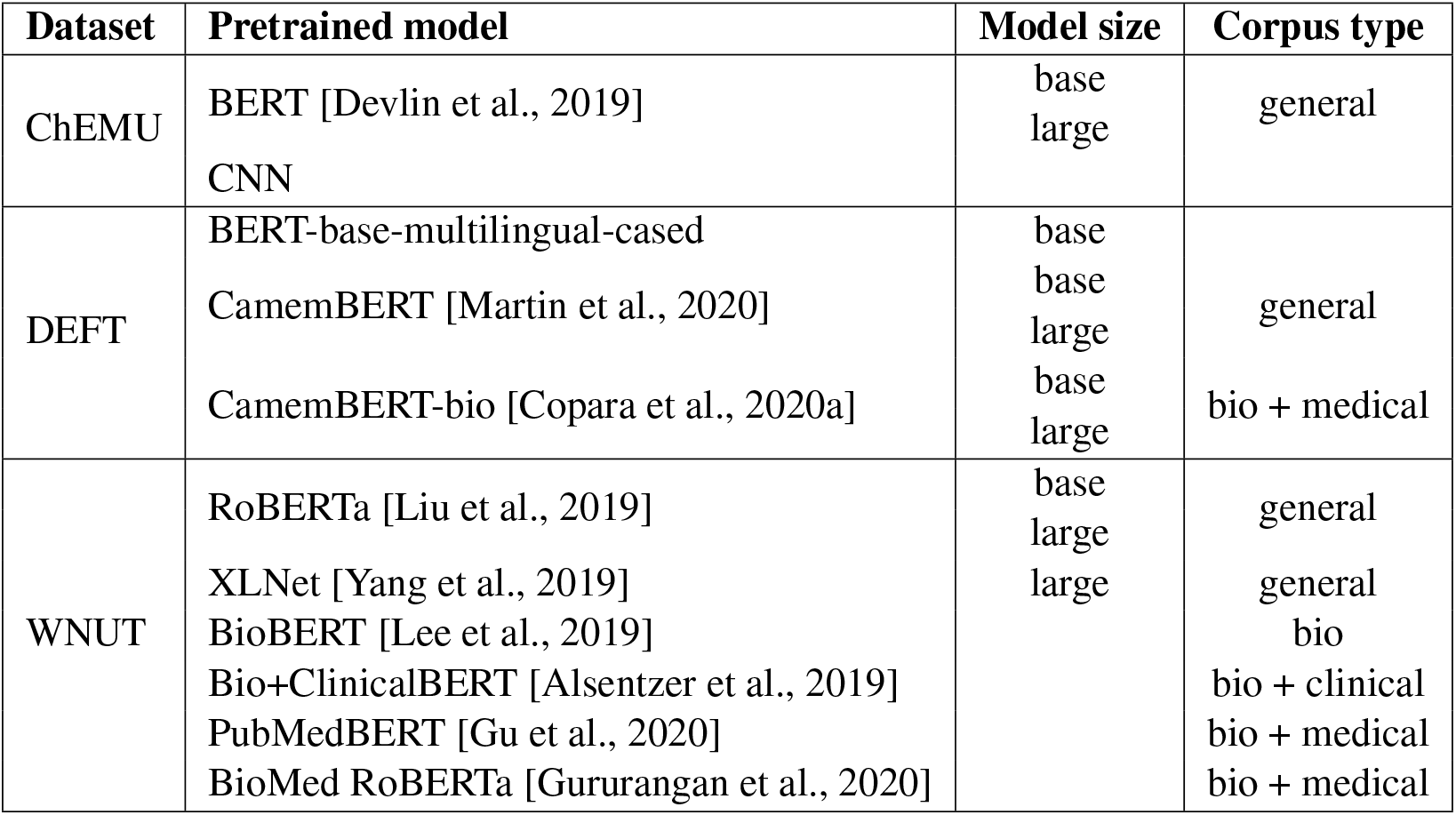
Pretrained models used for NER in the ChEMU, DEFT, and WNUT benchmark datasets.

To fine-tune a particular masked language model for the NER task, we leverage the token representation created in its pre-training phase. A fully connected layer is added on top of the token representations and trained to classify whether a token belongs to a class or not. As transformers usually use tokenizers that work on word bits (or sub-tokens), during prediction, the highest probable entity label will be assigned to all sub-tokens of a word and the sub-tokens will be then merged to build back the original word with the respective assigned label. Finally, in a given sequence, if two adjacent words were given the same entity prediction, we would consider the two words as a phrase related to that entity.

Following this approach, the masked language model is then fine-tuned on the domain-specific data - chemical, clinical and wet lab - using the training datasets previously discussed (ChEMU, DEFT, and WNUT). The fine-tuning is performed with the maximum sequence length of 265 tokens. The only preprocessing done was sentence-splitting. For the chemical and wet lab NER experiments, for which no nested entities were considered, we used a softmax function. Conversely, for the clinical NER, for which a token could be assigned to more than one entity, we used a sigmoid function to provide a multi-class classifier. More information about the fine-tuning of the models and the hyper-parameter settings can be found in [Copara et al., 2020b,a, Knafou et al., 2020].

### 4.2 Ensemble of deep masked language models for NER

Our ensemble method is based on a voting strategy, where each model votes with its predictions and a simple majority of votes is necessary to assign the predictions [Copara et al., 2020b,a, Knafou et al., 2020]. In other words, for a given document, our models infer their predictions independently for each entity. Then, a set of passages (token or phrases) that received at least a vote for the named entities is taken into consideration for casting votes. This means that, for a given document and a given entity, we end up with multiple passages associated with a number of votes, then, again for a given entity, the ensemble method will assign labels to all the passages that get the majority of votes. Note that each entity is predicted independently and that the voting strategy allows a passage to be labeled as positive for multiple entities at once, in case of nested entities. The individual models used in the ensemble models for the ChEMU, DEFT, and WNUT datasets are presented in Table 4.

### 4.3 Training and evaluation procedures

To train our models, we used the training subsets of the ChEMU, DEFT and WNUT. As shown in Table 5, we split this subset into train, dev and test sets to train the model weights, the hyper-parameters, and the best ensemble configuration, respectively. The ensemble threshold for chemical and clinical NER were set to 3 and, for wet lab NER, to 4. Then, a blind test set, which was provided as part of the official evaluation for the respective challenges, was used to evaluate the models. Table 5 shows the distribution of the splits for the different collections.

**Table 5:** Distribution of samples in the train, dev and test collections for the different NER tasks. Train: collection used to train model parameters. Dev: collection used to tune model hyperparameters. Test: collection used to define the ensemble models. Blind test: collection used to evaluate models.

| Dataset | Split | # patent snippets | # EHR notes | # wet lab protocols |
| --- | --- | --- | --- | --- |
| Training | Train | 800 | 80 | 370 |
| Training | Dev | 100 | 10 | 123 |
| Training | Test | 225 | 10 | 123 |
| Test | Blind test | 375 | 67 | 111 |

Results are reported in terms of the the micro and macro F_1_-score metrics and were computed using the BRAT eval tool^4^ against the blind test set split. The ensemble models created for the different domains are compared to a baseline based on BERT. Student’s t-test is used to assess the significance of the results. Results are considered statistically significant for p-values smaller than 0.05.

## 5 Results

In this section, we present the performance assessment of BERT as a NER baseline and the ensemble model over the test collection with the parameters identified in the training phase for the chemical, clinical and wet lab corpora.

### 5.1 Chemical NER results

Table 6 shows the performance of the baseline BERT and our ensemble models for the chemical NER using the F_1_-score metric in terms of exact and relaxed span matching. In the exact match evaluation, both the starting and the end offsets of the text spans of the predicted and gold standard reference entities match, whereas in the relaxed-match evaluation, the text spans of two entities overlap. The ensemble model achieves 92.30% of exact micro F_1_-score, yielding 1.3 percentage point improvement over the baseline (BERT-base-cased) (*p* = 0.005). It outperforms the BERT model for all the entities in exact match. If we consider the relaxed metric, the difference between the individual and ensemble model is minimal and not statistically significant (*p* = 0.07). These results indicate that the individual model is able to detect the passages containing the entities (or part of them) while the ensemble, by combining the power of different models, is able to predict the exact spans of the entities.

**Table 6:** Test phase results using the F_1_-score metric for the chemical patents NER (ChEMU dataset). BERT: BERT base-cased model. Ensemble threshold set to 3.

| Entity | BERT |  | Ensemble |  |
| --- | --- | --- | --- | --- |
|  | exact | relaxed | exact | relaxed |
| example_label | 96.17 | 97.30 | **96.69** | **97.84** |
| other_compound | 87.80 | 96.08 | **89.20** | **96.53** |
| reaction_product | 85.93 | **93.78** | **87.66** | 93.22 |
| reagent_catalyst | 87.91 | 90.82 | **90.22** | **91.76** |
| solvent | 94.44 | 94.91 | **95.41** | **95.41** |
| starting_material | 84.13 | 93.43 | **87.01** | **93.94** |
| temperature | 96.92 | 99.02 | **97.29** | 98.77 |
| time | 98.68 | 99.67 | **98.79** | **99.78** |
| yield_other | 97.99 | 98.21 | **98.42** | **98.65** |
| yield_percent | 99.36 | 99.62 | **99.74** | **99.74** |
| **all (micro)** | 90.98 | 95.96 | **92.30** | **96.24** |
| **all (macro)** | 92.93 | 96.28 | **94.04** | **96.56** |

The top-5 best performing entities identified by our models are *example_label, temperature, time, yield_other, yield_percent*. The entity with the lowest performance is *starting_material*, achieving 84.13% and 87.01% of exact F_1_-score for the baseline and ensemble models, respectively. Overall, the language models were effective to recognised chemical entities in patents, with a lower limit performance of 90.98% of micro F_1_-score for the single BERT model and as high as 96.56% of macro F_1_-score for the ensemble model.

### 5.2 Clinical NER results

The performance of BERT and the ensemble model for the clinical NER is summarised in Table 7. The ensemble model achieves 72.62% of overall micro *F*_1_-score, outperforming the baseline model by 8.82 percentage point (*p <* 0.001). Similar to the results obtained in the training set, the highest F_1_-score in the blind test set is achieved for the *valeur* entity (85.61%). This entity represents 7.6% of the annotations in the training collection. One could assume that entities with annotation examples above this threshold would perform well. However, when looking at the results for the *examen* (14.6% of the annotations) and *substance* (13.0% of the annotations) categories, we notice an important drop in performance (64.42% and 63.79%). Thus, it seems that the number of training data examples alone is not sufficient to learn an entity automatically.

**Table 7:** Test phase results using the F_1_-score metric for the clinical notes NER (DEFT dataset). BERT: BERT base-multilingual-cased model. Ensemble threshold set to 3.

| Entity | BERT | Ensemble |
| --- | --- | --- |
| anatomie | 76.46 | **80.69** |
| dose | 36.04 | **52.17** |
| examen | 68.42 | **73.33** |
| mode | 59.35 | **64.86** |
| moment | 67.48 | **78.69** |
| pathologie | 36.28 | **56.44** |
| sosy | 55.74 | **67.33** |
| substance | 55.86 | **63.79** |
| traitement | 45.98 | **60.76** |
| valeur | 81.60 | **85.61** |
| **all** (micro) | 63.80 | **72.62** |
| **all** (macro) | 58.32 | **68.37** |

The lowest performance for the ensemble method is found for *dose* entity. This can be due to the variety of values in the annotated data, combining numbers and words (e.g., *de 0,5 à 0,75 litre*), measure units (e.g., *1mg/kg/j*) or simply words that could be easily associated with a non-entity word (e.g., *24 paquets/année* or *02*). *Mode* entities are mostly words without abbreviations or numbers (e.g., *‘voie parasternale droite’, ‘voie centrale intraveineuse’*). Hence, they contain less variety for their values, which could lead to an easier way for the NER models to learn their patterns and make correct predictions. Nevertheless, results for the *Mode* categories are close to the median (64.86%). The lack of examples in the training set (3.2% of the annotations) could have impacted its performance negatively.

### 5.3 Wet lab NER results

The performance of BERT and the ensemble model for the wet lab NER is summarised in Table 8. As for the other domains, the ensemble model outperform BERT for all entities (*p* = 0.003), achieving an overall micro F_1_-score of 81.67%. The gain in performance is more relevant for the macro F_1_-score metrics, for which there is an increase of 3.28 percentage point between the baseline model and the ensemble model. In this case, it seems that the diversity brought by the ensemble enables the correct detection of more entity types.

**Table 8:** Test phase results using the F_1_-score metric for the wet lab protocols NER (WNUT dataset). BERT: Bio+Clinical BERT. Ensemble threshold set to 4.

| Entity | BERT | Ensemble |
| --- | --- | --- |
| Action | 87.35 | **88.35** |
| Amount | 84.47 | **86.76** |
| Concentration | 85.45 | **90.34** |
| Device | 69.66 | **71.46** |
| Generic-Measure | 40.00 | **50.51** |
| Location | 78.92 | **80.49** |
| Measure-Type | 68.61 | **68.07** |
| Mention | 70.10 | **73.09** |
| Method | 60.05 | **57.95** |
| Modifier | 55.82 | **58.11** |
| Numerical | 51.03 | **50.12** |
| Reagent | 89.19 | **92.03** |
| Seal | 86.84 | **90.13** |
| Size | 28.84 | **32.03** |
| Speed | 89.21 | **89.77** |
| Temperature | 90.12 | **94.15** |
| Time | 92.64 | **94.28** |
| pH | 94.74 | **95.31** |
| **all (micro)** | 79.34 | **81.67** |
| **all (macro)** | 74.71 | **77.99** |

Surprisingly, the entity with the highest F_1_-score, *pH* (95.31%), has only 0.2% of the annotations in the training sample. Again, the number of examples is not associate with the performance on the test set. Indeed, the best performing entities for the wet lab NER - *Temperature, Time* and *pH* - are responsible together for only 8.5% of the annotation examples. The lowest performance was found for the *Size* entity (32.03%), more than 2-folds worse than the average wet lab entity (77.99% for the ensemble model). The performance of the models are also low for the *Generic-Measure* and *Numerical* entities. Both have a small number of annotations in the training set (1.0% and 1.7% respectively). The *Generic-Measure* entity is similar to *dose* in clinical NER task and get various forms, such as measure units (*volume*), measurements (*30 kDa, 2*.*5 bars*, ∼*250–500 bp*), and ratios (*1:2, 1/500 to 1/1000*), which could also justify its low score.

## 6 Discussion

We compared the effectiveness of individual masked deep language models and ensemble models for the NER task for multiple domains and languages. Overall, the ensemble model improves the baseline BERT model by 4.6 percentage point, achieving an overall macro F_1_-score of 79.20% across all entities in the chemical, clinical and wet lab corpora. The ensemble model was relatively effective to recognised named entities in those corpora. Out of the 38 entity classes assessed, 45% had an F_1_-score higher 90% for the ensemble model compare to 24% for the individual BERT model. However, we notice an important performance reduction in the French corpus compared to the English corpora. This is likely due to the known issue of reduced resources compared to English, both in terms of the corpora to pre-train the masked language models but also to fine-tune for the clinical NER.

Figure 4 shows the comparison of BERT and the ensemble model for each dataset. The performance of the models on the clinical corpus is lower than on the chemical and wet lab corpora. This can be due to a smaller dataset available for fine-tuning the models. As seen in entity distribution tables (Tables 1, 2 and 3), the training data for chemical NER and wet lab NER are larger, which results in better performance of both the ensemble models and the BERT baseline. Additionally, the clinical dataset includes nested entities, which are known to be recognized more effectively using graph-based models [Yu et al., 2020]. Nevertheless, it is for the clinical dataset that we notice the highest relative gain in performance (14%) for the ensemble model.

**Figure 4:**
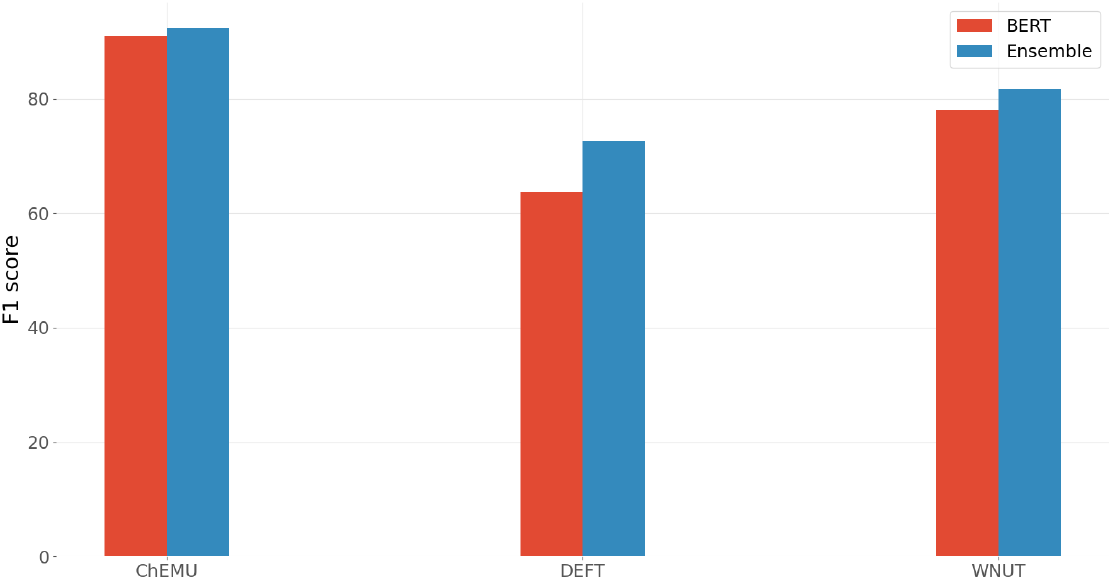
The performance of the ensemble model *vs*. BERT for different domains. Micro F_1_-score is used. The exact match is used for ChEMU dataset.

Our results show that the models have often difficulties recognizing infrequent entities, such as *dose* (clinical) and *Generic-Measure* (wet lab), which is inline with previous work [Fu et al., 2020]. However, we notice that for some entities, particularly in the wet lab corpora, the highest score were provided by infrequent entities. Indeed, as shown by Fu et al. [2020], a single holistic measure of F_1_-score cannot tell the details of performance of different models. Diverse entity attributes, such as *length, frequency, sentence length*, and *out-of-vocabulary (OOV) density*, are important for further model analyses.

To better understand our results across the different corpora, we performed a deeper analysis of the baseline and ensemble models using different entity properties: frequency, length, and label consistency. Figure 5 shows the comparison of the BERT baseline and ensemble models based on the frequency of the entities. For some infrequent entities, the ensemble model improves more over the BERT baseline. However, we also see performance gain for the ensemble model for frequent entities, such as *starting_material* (performance improvement of +2.9 point using the ensemble model). *starting_material* covers about 11% of ChEMU dataset and is more frequent than entities, such as *time* and *yield_percent*, for which the performance gains of the ensemble model are +0.11 point and +0.38 point, respectively.

**Figure 5:**
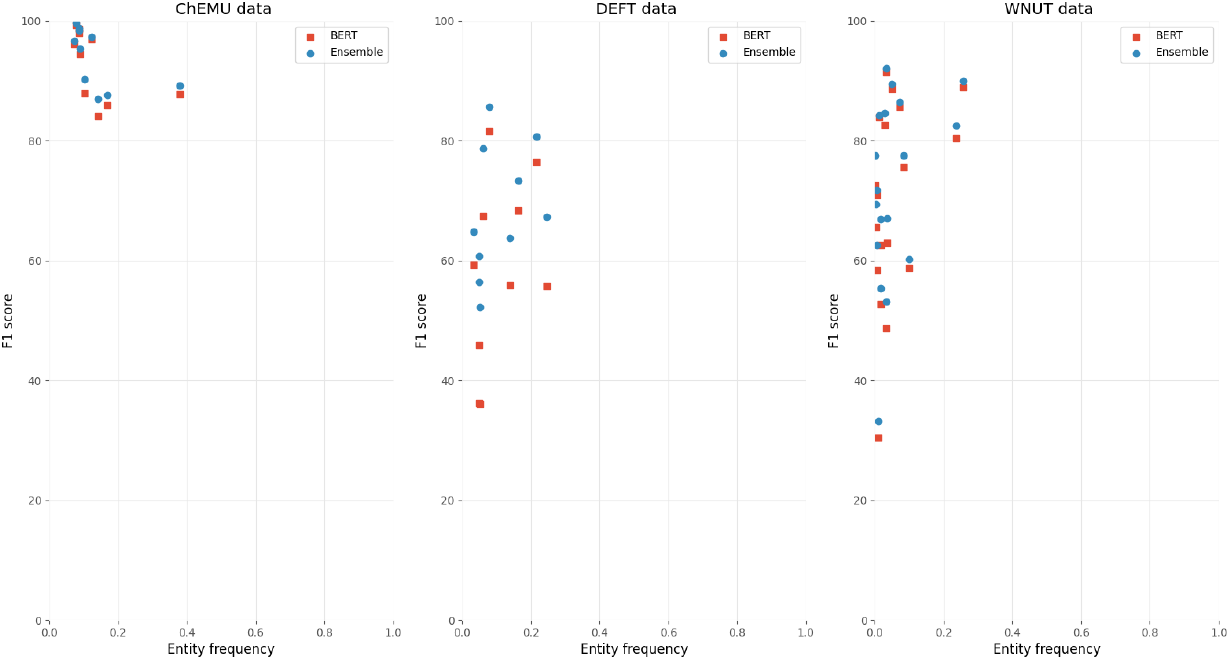
Performance of the BERT model *vs*. the ensemble model based on the entity frequency on the training data. For the clinical NER, the exact match scores are used.

Figure 6 shows the comparison of BERT and the ensemble model based on the entity length. The average entity length is shorter in the WNUT dataset. The ChEMU dataset, as expected, includes the longest average entity lengths. In general in all datasets, as the entity length increases, the performance of the ensemble model improves over BERT. In the ChEMU data, there is a direct link between the length and the performance of the models: the shorter the entity, the higher the performance.

**Figure 6:**
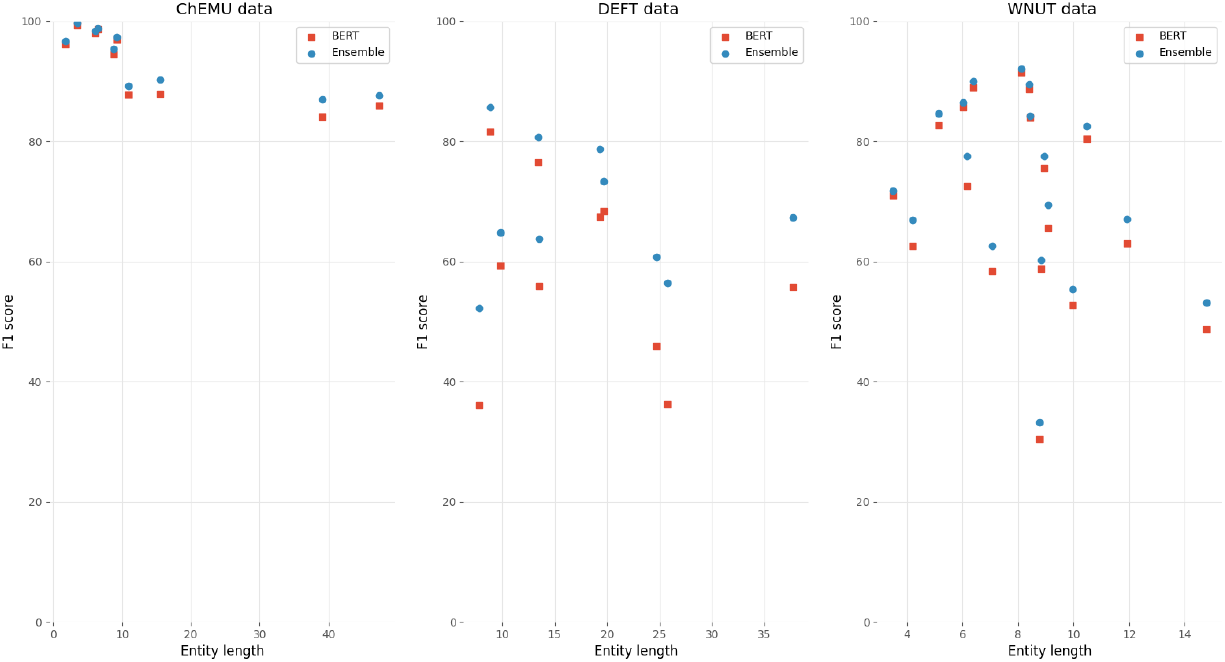
Performance of the BERT model *vs*. the ensemble model based on the entity length on the training data. For the clinical NER, the exact match scores are used.

Finally, Figure 7 shows the frequency of passages that were assigned more than one label for the three evaluated datasets. Here, we consider “passage” as a token or a sequence of tokens that were assigned a label, for example “triethylamine” annotated as *reagent_catalyst* and *other_compound* and “sodium hydrogen carbonate” annotated as *reagent_catalyst* and *other_compound* in ChEMU dataset, each was considered as one instance. The ChEMU and WNUT annotated corpora include passages that were assigned more than 2 labels, which makes them more ambiguous for the models. This can also explain the more modest performance of the models on WNUT dataset compared to the ChEMU dataset despite its larger data size.

**Figure 7:**
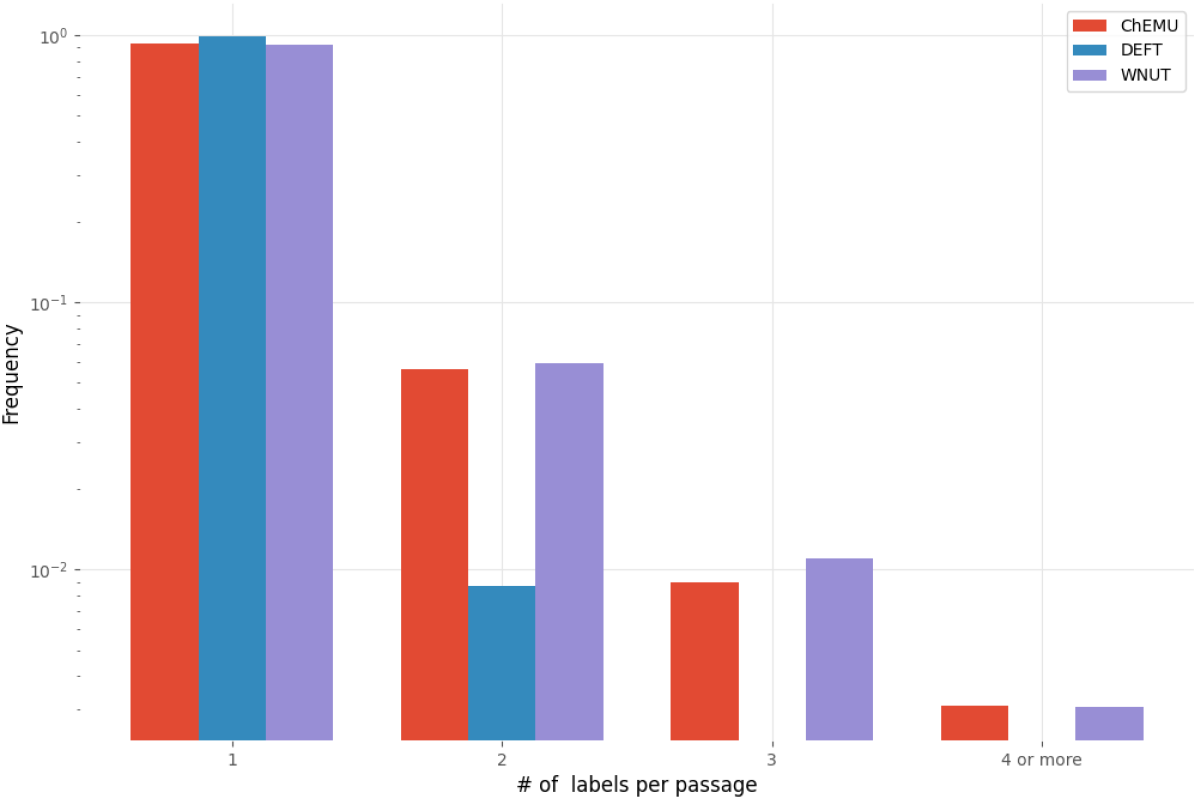
The number of labels assigned to each passage for the training set of the three datasets (ChEMU, DEFT, WNUT).

## 7 Conclusion

In this work, we compared the performance of the BERT model and its siblings for name entity recognition for three domains of chemical, clinical and wet lab, and English, and French languages. In all the domains and languages that we analyzed for NER, we show a significant improvement of performance by combining different deep masked language models compared to a strong baseline based on BERT single models. Overall, the ensemble model outperformed the baseline by 4.6 percentage point (*p <* 0.001), having 45% of the 38 entities assessed across the domains with a F_1_-score of 90% or more. We further performed a detailed analysis of the performance of the models based on a set of entity properties. We found that ensemble models can be more beneficial for longer entities.

## Data Availability

The data used for chemical NER can be found at: http://chemu2020.eng.
unimelb.edu.au/. The data used for clinical NER can be found at: https:
//deft.limsi.fr/2020/. The data used for wet lab NER can be found at:
http://noisy-text.github.io/2020/wlp-task.html.

## Author Contributions

NN drafted the manuscript, implemented the models and analysed the results. JK designed and implemented the models, and analysed the results. JC implemented the models and analysed the results. PR analysed the results. DT drafted the manuscript and analysed the results. All authors reviewed and contributed to the writing.

## Data Availability Statement

The data used for chemical NER can be found at: http://chemu2020.eng.unimelb.edu.au/. The data used for clinical NER can be found at: https://deft.limsi.fr/2020/. The data used for wet lab NER can be found at: http://noisy-text.github.io/2020/wlp-task.html.

## Footnotes

1 https://data.mendeley.com/datasets/wy6745bjfj/2

2 https://jep-taln2020.loria.fr/ateliers/deft/

3 https://www.protocols.io/

4 https://bitbucket.org/nicta_biomed/brateval/

